# Towards Development of Guidelines for Virtual Administration of Standardized Language and Literacy Assessments: Considerations for Clinicians and Researchers

**DOI:** 10.1101/2021.06.07.21258378

**Authors:** Emily Wood, Insiya Bhalloo, Brittany McCaig, Cristina Feraru, Monika Molnar

## Abstract

**Objectives:** Previous virtual care literature within the field of speech-language pathology has focused primarily on validating the virtual use of intervention programs. There are fewer articles addressing the validity of conducting standardized paediatric oral language and literacy assessments virtually. Additionally, there is a lack of practical recommendations available on how best to conduct these assessment measures virtually. Given the rapid rise in virtual care as a result of the COVID-19 pandemic, clinicians and researchers require guidance on best practices for virtual administration of these tools imminently.

**Methods:** We identified six key themes for recommendations based on two sources (1) our lab meeting discussions and (2) the outcomes of a narrative review of the extant literature. We then conducted semi-structured interviews with a group of 12 clinicians, students and researchers who had administered standardized language and literacy assessments with a variety of monolingual and multilingual school-aged children, with and without speech and language difficulties, in clinical and research settings to generate recommendations within these themes. Subsequently, in line with the Guidelines International Network, these recommendations were rated by group members, and reviewed by external stakeholders. A quasi-delphi consensus procedure was used to reach agreement on ratings for recommendations.

**Results:** We have developed recommendations for the use of standardized language and literacy assessments in virtual care, across six key themes: candidacy for virtual assessment, communication and collaboration with caregivers, technology and equipment, virtual administration, ethics, consent and confidentiality, and considerations for bilingual populations.

**Conclusions:** This paper is one of the first to share practical recommendations for virtual assessment in the domain of paediatric oral language and literacy assessment. We hope these recommendations will facilitate future clinical research and that as the body of research grows, that this paper will act as a basis for the development of formal Clinical Practice Guidelines.

## Introduction

Virtual care, formerly or alternatively known as telepractice, telehealth, remote care, or telemedicine, is any interaction between patient or client and a member of their circle of care, occurring remotely, using any form of communication or information technologies with the aim of facilitating or maximizing the quality and effectiveness of patient care.^1^ There is no consistent and universally agreed-upon term for this service delivery model. For the purposes of this paper, we will use the term most often used in Ontario, virtual care, as this is where these recommendations were developed.

The use of virtual care has been steadily growing, but saw a recent surge in 2020, in large part due to restrictions imposed by the Coronavirus-19 (COVID-19) pandemic, which obligated many health care professionals and researchers to begin working remotely. In the United States, primary care virtual visits increased by 50% from 2019 to 2020.^2^ Similarly, Canada Health Infoway reports that in so far 2021 40% of primary care visits were conducted virtually, approximately double the tele-visits pre-pandemic.^3^

Virtual care is advantageous for various reasons: clinicians can save time and provide care to a greater number of individuals,^4^ clients who live in rural areas without clinics or those without transportation can access clinical services remotely,^5^ those with physical disabilities that make travelling challenging, or those who feel more comfortable undergoing assessment in their own homes, also benefit from virtual care.^6^ Furthermore, bilingual clients seeking clinicians who speak their language(s), and those seeking practitioners with knowledge in a specific area of practice may be more likely to find a suitable match when they can search a broader geographical area.^7^ This is especially important when we consider that although approximately half of the world’s population is bilingual,^8, 9^ only 6.5% of ASHA registered Speech-Language Pathologists (S-LPs) report being bilingual,^10^ and 73% of Canadian S-LPs report there being a lack of availability of other S-LPs who speak the language of their diverse client populations.^11^

Despite these many advantages, a 2011 ASHA survey reported that only 2.3% of delivery of any Speech Language Pathology (S-LP) service was conducted via virtual care, and only 11% of clinicians had used virtual care.^12^ This earlier reluctance to embrace virtual care could have been a result of perceived barriers to the provision of this type of service. In a 2016 paper, Sutherland provides a review of these barriers reported by clinicians in Australia.^13^ Three categories of barriers were identified: financial, technological, and cultural. Many clinicians felt that the upfront costs of providing virtual services were prohibitive, both for the clinicians and the communities they serve. They also reported that there was limited access to, training in and familiarity with technology for clinicians and clients and that addressing this issue would require significant support from policy makers and government officials. Additionally, there were concerns on behalf of clinicians that rural families would not embrace or be interested in accessing services virtually. This notion was not supported by research, which in the contrary suggested these families were interested in this service delivery model, largely in part due to the lack of face-to-face clinics available in their communities. ^14^ Many clinicians, particularly those in urban areas, indicated that virtual care was a service that would be uniquely valuable to those in remote areas, and were minimally interested in pursuing this model.

Evidently, the widespread integration of virtual care accelerated in Spring 2020, with the intensification of the COVID-19 pandemic. In 2016, StatsCan reported that 4% of the population worked from home, while 32% still work remotely now in 2021. ^15^ Clinicians and researchers adjusted and modified their work as schools, clinics and laboratories began operating virtually during the pandemic, demonstrating that many of these barriers were often surmountable when no other options were available. In fact, a recent study from Sutherland in May 2021, collected feedback in the form of a questionnaire from 27 pediatric S-LPs, who conducted CELF-5 assessments virtually. Their qualitative reports suggested that the primary obstacle was overcoming technical issues in virtual assessment, but that otherwise virtual assessments were easy to conduct, and still allowed for connection with clients throughout the pandemic.^16^ Now that working remotely is so commonplace, it is likely that virtual clinical assessment and research techniques will remain a trend in the future, especially considering that in 2021, a reported 82% of U.S. Employees, and 80% of Canadian employees indicated interest in working from home at least some of the time after the pandemic ends.^15^

Owing to the fact that virtual care is a relatively new service delivery model in both clinical work and research, there is still a subsequent paucity of literature related to its application.^17^ Existing studies suggest that virtual care is a feasible, effective, and appropriate alternative or addition to face-to-face practice for S-LP clinicians and researchers.^17^ Thus far, many studies have focused on the implementation of specific treatment programs or therapy methods through virtual means.^17–22^ Several studies have validated the use of specific treatment programs for adults and children in a virtual service delivery model across domains of practice. This includes but is not limited to instruction in reading and spelling for children,^23^ speech sound intervention in children ages 6-10 ^24^, stuttering treatment for young children using the Lidcombe Program ^25^, and for adolescents using the Camperdown Program ^26^, voice intervention for adult patients with Parkinson’s using the Lee Silverman Voice Treatment LOUD intervention, ^27^ and in the management of dysphagia in adults. ^28^ In contrast, fewer studies have examined the validity and feasibility of conducting assessments, specifically standardized assessments online. ^29^ This is interesting, in part because while the ultimate goal of intervention is the functional use of language in real life scenarios, this contextual support could be more challenging to promote in a virtual care setting, where clinicians and clients are experiencing different circumstances.

Standardized assessments, however, aim to evaluate the building blocks of skills like language and literacy in the absence of this contextual support. These assessments are designed to evaluate specific language competencies in isolation. Consequently, it is possible that standardized assessments may be well-suited to virtual administration.

Of the few studies pertaining to standardized assessment, most have focused on the validity and virtual administration of standardized assessment tools in the adult population, rather than the pediatric population. These studies have primarily focused on validity of assessments for virtual use and indicate high rates of agreement between online and in-person evaluation for standardized assessment of neurogenic communication disorders using the Boston Diagnostic Aphasia Examination in post-stroke adult patients,^30^ non-standardized evaluation of oral motor skills, swallowing and communication ability in post-laryngectomy adult patients,^31^ and standardized assessment of linguistic skills in post-stroke adults using the standardized instrument, the Western Aphasia Battery.^32^ There have been few studies that have evaluated validity of commonly used oral language and literacy assessments for virtual use in the pediatric school-aged population.^29^

Of the published studies exploring validation of virtual use of standardized language and literacy assessments, there is some evidence to suggest that certain subtests of the Clinical Evaluation of Language Fundamentals, are valid (CELF-4) ^33^ and amenable (CELF-5) ^16^ for use in a virtual setting. Additionally, the makers of the Test of Integrated Language and Literacy Skills (TILLS), conducted a study in 2020 which reportedly validates their product for virtual use, as Tele-TILLS ^34^. However, this validation paper is not made available on their site.

Furthermore, many of these validation studies are based on research conducted or funded by the test makers themselves. These groups may have an inherent bias to validate their tools for virtual use, when this has been one of the only available models of service delivery throughout the pandemic. Regrettably, few other in-person tests are validated for virtual use. In response, many test makers have created online or virtual versions of their most common tests and recommend use of these products in a virtual setting. For example, Pearson indicates that the online Q-global version of the CELF-5 can be used reliably in a virtual setting.^35^ However, many clinicians and researchers may not yet have access to or training in the use of these virtual materials. To address this issue, some test makers like Pearson,^36^ which produces oral language measures such as the Expressive Vocabulary Test [EVT],^37^ and Pro-Ed,^38^ which develops the Comprehensive Test of Phonological Processing [CTOPP],^39^ have issued No Objection orders, allowing S-LPs and researchers to use portions of their test materials virtually through non-public facing teleconference software, provided they follow their rules for administration, and in certain occasions, ask for their permission.^35^

Despite the limited number of independent studies validating virtual standardized assessment for tests of oral language and literacy, regulatory bodies such as Speech-Language Pathology and Audiology Canada [SAC],^40^ ASHA,^6^ Royal College of Speech Language Therapists,^41^ Speech Pathology Australia,^42^ and the Indian Speech and Hearing Association, ^43^ have authorized clinicians to proceed with assessment of these skills via virtual care. They are cognizant that as a result of the COVID-19 pandemic, clinicians are in unique position where are required to continue to provide accurate, timely and informative assessment through virtual care, while research into the validation of their assessment materials is ongoing. Permission to continue these evaluations is critical, as strong oral language skills and proficient literacy abilities have both been well-established as skills that are associated with later positive life outcomes, such as adolescent social and emotional skills and adult socio-economic status and acquired education level. ^44, 45^ In their work, researchers and clinicians commonly use standardized assessment measures to assess, identify difficulties, and provide early intervention for mitigating potential oral language or literacy difficulties.

Oral language comprises knowledge of five areas: *phonology* (the system of speech sounds in a language); *morphology* (the smallest meaningful units of language and how they are combined); *semantics* (the understanding and use of words and phrases); *syntax* (the rules that governs how words and phrases can be combined); and *pragmatics* (the social norms that dictate how language is used with others in context).^46^ S-LPs have extensive knowledge of these domains and play a vital role in the evaluation and management of all aspects of oral language, from birth to adulthood. This role often includes the use of standardized tests. ^47^ Because of their knowledge of oral language and its relationship with reading and writing, S-LPs are also regularly involved in the assessment and management of literacy difficulties in children as well. ^48^ Literacy is defined as “the ability to identify, understand, interpret, create, communicate, and compute using printed and written materials associated with varying contexts”.^49^ It is broken down into three main skills, reading; the process of converting symbols in print into identifiable words, which includes the ability to decode and to comprehend writing; the process of expressing thoughts and ideas using printed symbols to make up individual words and sentences, and spelling; the ability to separate words into their individual sounds, and link those sounds with specific letter or letter combination to create a word.^48^ S-LPs assess and treat discourse level skills like reading comprehension and written expression, as well as word-level skills like phonological awareness or the ability to hear and manipulate parts in words. ^48^ Again, the role of the S-LP in literacy often includes using standardized tools to evaluate these component skills.

Given that standardized oral language and literacy assessments are often a critical piece of a comprehensive speech and language assessment from both clinical and research perspectives, ^50–52^ and those using these tools may be required to conduct these assessments virtually, clinicians and researchers require to up-to-date guidelines to inform their virtual practice, so that they can continue to provide these valuable standardized assessments.

Standardized tests have historically been created for use in in-person settings, and many components of the virtual testing process may be different or unfamiliar to clinicians and researchers who are used to in-person administration but have had to adapt their practice to current circumstances. Practice documents from regulatory bodies like CASLPO and SAC, as well as guideline papers published in academic journals,^30^ have focused primarily on technical, licensure and ethical considerations, and less on the clinical and practical components of assessment, such as the role of the caregiver, organization of the testing space or manipulation of testing materials.^5, 40, 53^ Documents from other international regulatory bodies, like the Telepractice page available on the ASHA website, provide practical resources dedicated to best practice in virtual care (telepractice), including a section specific to assessment. However, this assessment page only briefly discusses test modification, standardization, and documentation of results, and does not address other topics like the role of the caregiver in assessment or specific administration recommendations. ^54^ In addition, the process for developing this ASHA assessment page is not described or available for review. A brief list of references is provided, but no information is given regarding the authors of the document, their qualifications, their experience with virtual care or how they decided what recommendations to include or exclude.

### Scope of the Recommendations

The objective of this paper is to provide recommendations for using commonly assessed standardized tools in a virtual setting. These include receptive and expressive skill evaluations within the domain of oral-language and cognitive-linguistic skills as well as reading, writing, and spelling. These recommendations are intended for clinicians and researchers who use standardized language and literacy assessments, within these domains, in the context of virtual care or research. In order to address the identified gap in the literature, the recommendations presented here consider the practical components of assessment – as opposed to technical considerations typically addressed by regulatory bodies and validation studies. They explore themes commonly discussed in the existing virtual care literature, as well as those that pertain to issues raised by group members engaging in virtual administration of assessments. The themes include candidacy for virtual assessment,^17^ technology and equipment management,^55^ virtual administration of test protocols,^56^ ethics, consent, and confidentiality,^56^ in addition to communication and collaboration with caregivers and considerations for bilingual populations – the latter two themes emerged based on the current study.

When considering previously published guidelines, these recommendations are unique and address gaps in the literature. We provide a resource that is useful for both clinicians and researchers, that is based on the best current available evidence, as well as lived clinical and research experience conducting assessments virtually. Furthermore, the development of these recommendations is reported in a transparent way and is replicable if needed. Lastly, we have no bias to recommend the use of specific products or assessment tools in our paper. Note: A preprint of this paper has been uploaded to the medRxiv server.^57^

## Methods

### Ethics Statement

This study has been partially funded by the University of Toronto’s COVID-19 Student Engagement Award (RIS Human Protocol Number: 38608). No conflicts of interest, financial or otherwise, are declared by the authors or members of the guideline development group.

### Composition of “Recommendation Development Group”

The recommendation development group is comprised of 12 members, with varied experiences using virtual platforms to conduct standardized language and literacy assessment measures with school-aged children.

The initial group was comprised of 10 individuals conducting virtual assessments as part of a study examining online, as compared to in-person, assessments of standardized oral language and literacy tools in monolingual and bilingual children at the University of Toronto (Department of Speech-Language Pathology). It included a clinical speech-language pathologist with six years of experience working for a school board, one year of which was exclusively in a virtual service delivery model; a second-year research MSc student studying bilingual literacy development; two second-year clinical S-LP students; and 6 research assistants from the University of Toronto. This group conducted standardized assessments on a group of typically developing English-speaking monolingual (n=81) and linguistically diverse bilingual children (n=99), aged 4-8 years (n total=180). Typically developing children in this age group were assessed, rather than children with identified language and literacy disorders, as this was part of the inclusion criteria for this study. While clinicians do not regularly assess typically developing children, researchers often do, and this paper is meant to serve both clinicians and researchers.

Group members participating in this study, evaluated expressive vocabulary (via the Expressive Vocabulary Test-2; EVT-2 ^37^), phonological awareness (via the Comprehensive Test of Phonological Processing – 2; CTOPP-2 ^39^), and word and non-word reading (via the Woodcock Reading Mastery tests ^58^). All tests were conducted in English. This ongoing tele-assessment study commenced in April 2020, after the COVID-19 pandemic and is ongoing.

In order to gather further clinical insight in the process of developing the recommendations, two additional clinical S-LPs each with over 15 years of experience of face-to-face practice experience, were recruited for participation, for a total of three clinical S-LPs. These clinicians, worked in an exclusively virtual setting for the 2020-2021 school year. As part of their clinical work, they conducted virtual standardized assessments on school-aged children with speech and language difficulties, either for the purpose of formal identification in the school system, or for the provision of specific programming for educators and caregivers. These clinicians used a variety of standardized assessment tools, which included but were not limited to; the Clinical Evaluation of Language Fundamentals-5 (CELF-5) ^59^, the Test of Integrated Language and Literacy Skills (tele-TILLS) ^60^, the Test of Auditory Processing Skills-4 (TAPS-4) ^61^, the CUBED Narrative Language and Dynamic Decoding Measures ^62^ and the Expressive Vocabulary Test –3 (EVT-3) ^63^.

In summary, the final group consisted of 12 individuals with varying backgrounds and experiences administering standardized language and literacy assessments virtually. Several group members were novices, either new to research, like the research assistants, or new to clinical practice, like the S-LP students. Others were more experienced in their respective fields, like the M.Sc. student, or the school-board clinicians. Some had experience evaluating typically developing students, like those participating in the study, and others primarily assessed children with suspected or identified language and literacy difficulties, like the practicing clinicians. A few members had more practice using assessment tools virtually, while others had primarily conducted assessments face-to-face. All members, novice or expert, researcher, or clinician had experience conducting standardized assessments with school-aged monolingual and bilingual children. The diversity among the 12 group members ensure that a broad spectrum of experiences are reflected in this recommendation paper.

### Development of Recommendations

The following recommendations (see Results section) have been developed in accordance with the Guidelines International Network framework (G-I-N).^64^ These recommendations are intended to act as a step toward the development of Clinical Practice Guidelines for virtual standardized assessment, and as additional research becomes available, should be revised, and updated. These recommendations were developed between August 2020 – June 2021. See Figure 1 (below) for the development process.

**Figure 1.**
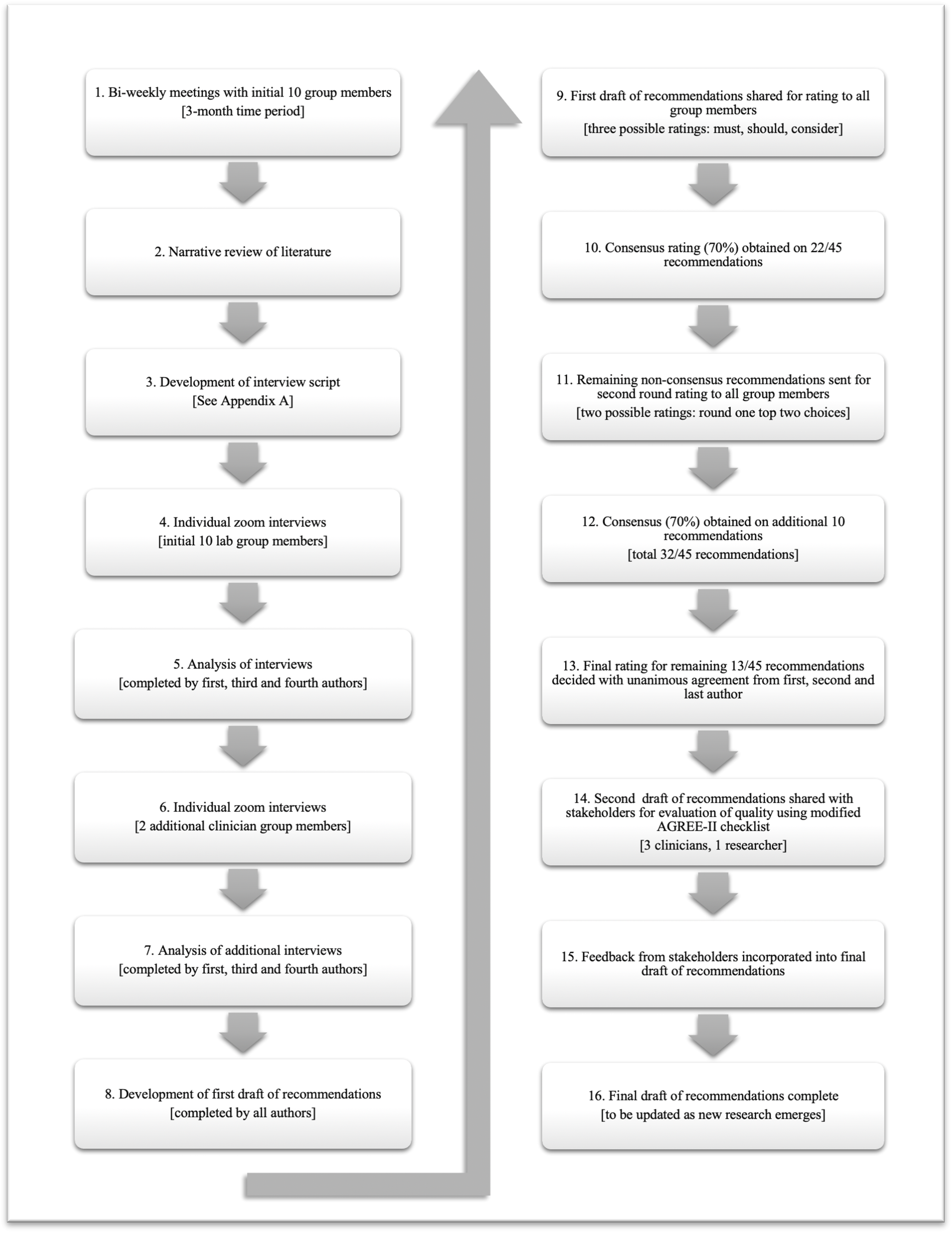
Process for Development of the Recommendations

### Team Meetings

While conducting virtual assessments as part of the previously mentioned online literacy study, the 10 initial lab group members met weekly, or bi-weekly, to discuss issues relating to test administration and to provide feedback and advice to one another. These conversations between lab members were unstructured, and a variety of topics, including behaviour management, communication with caregivers, online scheduling, connectivity and technology troubleshooting, and test administration rules were addressed. These problem-solving and trouble-shooting discussions served as a starting point for the development of an informal shared document that provided practical, useful information for administering standardized assessments virtually. The group members subsequently determined that this information would be valuable

for clinicians and researchers working in other settings beyond the lab, and as a result, the process of developing the current recommendation paper was initiated.

### Development of Key Themes

The themes addressed in this paper were developed from two sources. First, select themes were incorporated based on information from papers identified in a literature search, conducted in August-September 2020. For this narrative style review, two search concepts were used on MEDLINE and Embase databases: 1. Telemedicine (telepractice, telehealth, telemedicine, virtual care) and 2. Speech-Language Pathology (speech therapy, speech disorder). This search yielded a total of 176 articles. 129 of these were deemed irrelevant and removed, as they did not relate to language or literacy, along with 12 duplicates, leaving a total of 35 unique articles. Of these 35, 30 were excluded as they either; discussed intervention or treatment in language and literacy rather than assessment; were conducted on preschool or adult populations rather than with school-aged children; or were conducted face-to-face rather than virtually. Only five specifically addressed virtual language or literacy assessment and screening in a virtual setting with school-aged children, ^33, 65–68^ Three papers discussed issues pertaining to language assessment, and two examined validity and reliability of conducting literacy assessments online. See Appendix A, Table 1 for a summary of these articles.

Subsequently, the authors identified an additional three papers from this narrative review which provided general guidelines for virtual practice in Speech-Language Pathology.^17, 55, 56^ Review of these guideline articles identified common themes which were used to guide the development of the recommendations. See Appendix A, Table 2 for a summary of these guideline papers.

Importantly, this narrative review revealed a lack of evidence to inform clinical practice regarding virtual assessment. The small number of articles was not felt to be sufficient to warrant a systematic review of the evidence which would support the development of formal Clinical Practice Guidelines. Rather than wait for publication of additional papers on this topic, the authors decided to combine the findings of this narrative style review, with lived experiences of clinicians and researchers to create a set of recommendations that could be readily available during the COVID-19 pandemic.

Consequently, the themes outlined in this recommendation paper are primarily based on information from the three guideline papers identified in the narrative review, and the experience of the group members conducting assessments virtually. Specifically, the guideline papers lead to the identification of four themes — *administrative, technical, and ethical issues*,^55^ *clinical considerations*,^56^ *client candidacy,*^17^ and the team meetings with group members yielded two additional themes — *communication and collaboration with caregivers* and *considerations for bilingual children*. The combination of the narrative review and the team meetings yielded a total of six themes. These themes were derived from the best available scientific evidence, as well as the clinical expertise of the researchers and clinicians involved, both of which are described as critical components of evidence-based practice, according to ASHA’s position statement on Evidence-Based Practice in Communication Disorders.^69^ For a summary of the themes and the sources that inspired them, see Tables 2 and 3 in Appendix A.

### Individual Interviews

Subsequent to the identification of the six themes, an interview script was developed and administered by the first author with each group member (see Appendix B). The six themes, as previously stated, are: *Candidacy for Participation in Virtual Assessment, Communication and Collaboration with Caregivers, Technology and Equipment Considerations, Virtual Administration of Standardized Assessments, Ethics, Consent and Confidentiality, and Special Considerations for Bilingual Populations*. An additional section titled “*other*” was included in the interview script to address any outstanding points or issues that group members felt did not align with previously identified themes. Members were also encouraged to contact the first author by email after the interview if they had any additional points for consideration. The first author had a previously established working relationship with all group members, whether through volunteer research at the lab for the initial ten members, or through clinical work at the school board for the two clinical members. Group members were aware that they were participating in the development of recommendations for virtual standardized assessment and were familiar with the experience and the objectives of the first author.

First, group members from the lab with experience conducting standardized assessment virtually were purposefully selected for participation in the interview and development of the recommendations. Initially, members were asked as a group to participate virtually during a lab meeting, and following this general invitation, were sent a follow-up email by the first author confirming a time for their individual sessions. The interviews were conducted virtually over the course of a two-week period, and typically lasted 30-40 minutes. The first author acquired verbal consent to record the sessions, and all sessions were audio and video recorded on Zoom. No other persons were present at the time of the recording and all interviews were conducted from the first author’s home.

The first author used the interview script, which addressed the previously identified six themes, to guide the conversation. Group members were asked to frame their responses in terms of challenges and advantages associated with each theme and were given as much time as they needed. Interviews were semi-structured, and the group members were provided the opportunity to discuss the advantages and challenges associated with each theme, as well as potential solutions they may have identified, and problems that persisted. All recruited members participated in the interviews. Upon completion of the interviews, it was determined that additional clinical input from S-LPs working virtually outside the lab would help strengthen the recommendations and render them more clinically significant. The first author recruited two additional clinical S-LPs from a school board for participation in individual interviews. These clinicians were selected because they had extensive in-person clinical experience working with school-aged children, and they were both currently working virtually for a school board in the 2020-21 school year. They participated in the same interview process as the other group members. Each of the 12 members was interviewed only once and group members were not provided with a copy of these notes for review.

### Analyzing the Interviews and Compiling the Recommendations Draft

The first author listened to and took notes on all 12 interview audio files. Observations were recorded and noted in a table with a challenges and advantages for each of the six themes. Subsequently, the third and fourth authors each independently listened to and evaluated 6 recordings each and noted their observations in their own notes document. The first author then integrated all noted observations into one document. See Supplemental File 1 for the rough notes from these interviews. This methodology is consistent with a deductive style analysis (e.g.,,^70–71^), as themes were identified prior to conducting the interviews and based off narrative review of existing literature, and discussions in lab meetings. The authors elected to take this approach for several reasons. First, presenting each member with an opportunity to address all themes, ensured that every member had the chance to discuss and contribute equally to each section, and that there would be robust discussion across a wide variety of topics. Secondly, deductive analyses are useful when time and resources are limited. Given the rapid uptake of virtual service delivery as a result of the pandemic, the authors felt the need to make these recommendations readily available as soon as possible, and thus selected the most time-sensitive approach. In order to address some of the potential bias of structuring our interviews around themes, the authors elected to include an “*other*” discussion point at the end of each interview. This provided each group member the option to add additional points that may not have felt aligned with a specific, previously identified theme.

Subsequently, the first, third and fourth authors collectively developed the first set of recommendations from the aggregate of the interviews. No direct quotes from the group members were used in the recommendations. Rather, the authors identified key points from the interviews for inclusion in the recommendations, based on how often a topic was discussed and its’ general relevance. The authors elected to take this approach in order to be consistent with existing recommendation papers from published scientific studies and from regulatory bodies that typically present broad general statements or directives, rather than direct quotes from a specific clinicians or researchers.^40–42, 53–56^ There was a great deal of overlap in responses to each theme from all group members, and only few minor, or overly specific ideas. For example, nearly all group members mentioned the importance of a strong internet connection, and the need to prepare for and plan reinforcement and breaks in an online testing session, so these points were included as recommendations in the first draft. However, overly specific, or infrequently mentioned points, like relabelling the names of audio files associated with specific tests for ease of recognition, were not included. The authors collectively used these notes to draft the first set of the recommendations, which were grouped into the previously identified six themes for ease of use and readability. This resulted in a total of 45 recommendations across the six themes that were directly derived from the interviews.

### Decision-Making Process for Ratings

This initial draft of the recommendations was then disseminated to all 12 group members, without qualifying descriptive adjectives, via a survey to allow for their rating. Members were instructed to rate each guideline, by choosing either “must,” “should,” or “consider.” They were informed that the “must” rating referred to recommendations that are required/mandatory to be followed in order to comply with regulations from their regulatory governing bodies such as those from CASLPO. For example, the obligation to obtain informed consent for all aspects of virtual care, and the importance of determining whether a client is appropriate for this type of service delivery.^72^ The “should” rating referred to recommendations that should be followed whenever possible or feasible, to ensure best practice, but that are not necessarily mandated by a governing body. For example, suggesting the assessment take place in a quiet, separate room with a caregiver present is preferred, but not always possible for every family depending on their housing situation, and their other caregiver and work obligations. The “consider” rating referred to recommendations that are merely valuable pieces of additional information for consideration that clinicians and researchers might find useful as they begin their virtual assessments. For example, if available, using two devices to allow for better observation of the child while simultaneously displaying testing information.

A Quasi-Delphi method was used to survey group opinion regarding the recommendations, with decision-making consultations between the first and second authors, along with the principal investigator, at the first and second rounds of survey. Previous studies, adopting the Delphi method, recommended 2-4 survey rounds and a minimum of 50%-80% consensus at each survey round.^73^ Similarly, we aimed to conduct a minimum of 2 survey rounds, with a mid-level consensus threshold of 70% per round, until consensus was achieved. Recommendations were approved and adopted in instances where a minimum of 70% of group members agreed on their rating. Review of the first round of ratings indicated that the group members reached 70% consensus on 22/45 recommendations. Subsequently, a second version of the survey was shared with group members, after consultation with the first and second authors as well as the principal investigator, regarding unresolved recommendations that did not achieve group consensus. In this second survey version, members were asked to rate the remaining unresolved recommendations again. In this version, only the top two choices chosen in the first survey were provided as options. For example, if an unresolved recommendation had a 50:40:10 percent rating for “must”, “should” and “consider”, only the top-two rating options, “must” (50% consensus) and “should” (40% consensus), were sent to group members for re-rating. In order to ensure rating accuracy and avoid response bias, group members were not informed of agreement percentages per rating option as well as which of the top-two rating options had greater agreement. Review of the second survey ratings indicated the group had reached consensus on an additional 10 recommendations. All 12 group members participated in both rounds of recommendation rating. To decide on a final rating for the remaining recommendations, the first and second author, along with the principal investigator met to discuss and determine the final rating for the remaining 13/45 recommendations. The final rating was determined when the three group members unanimously agreed. A breakdown of the rating of votes for survey 1 and survey 2 can be found in the Appendix C. After finalizing the ratings, the recommendations were complete and were shared with community stakeholders for review and feedback.

### Peer Review and Stakeholder Consultations

In order to obtain feedback on the quality of the recommendations, a draft was shared with four stakeholders, three additional school board S-LPs working in a virtual care setting and one additional researcher who reported using standardized assessments tools virtually in their work. The two school board S-LPs each had more than five years of clinical experience, and a full year of experience working in an exclusively virtual setting at the time of the consultation. The researcher was a fourth-year student in a combined MClSc/PhD program with more than 3 years of lab experience, and recent direct experience conducting standardized assessments in a virtual setting with children. These external stakeholders were asked to evaluate the quality of the recommendations using an adapted version of the Appraisal of Guidelines for Research & Evaluation Instrument (AGREE-II).^74^ We adapted the AGREE-II checklist, developed for intervention-based clinical guidelines, as there are no checklists specific to assessment guidelines development (regardless of assessment medium, whether online or in-person). The AGREE II is comprised of 23 items sorted into 6 categories: Scope and Purpose, Stakeholder Involvement, Rigour of Development, Clarity of Presentation, Applicability, Editorial Independence.

Individual items are rated on a scale of 1-10 and the checklist is scored within each category using a percentage, with a score of 70% indicating a high-quality guideline. In our modified version of the AGREE II, we retained the 6 categories, but adapted select items within each to better reflect the practice to evidence based approach used in the development of these recommendations. We also opted to emulate the International Centre for Allied Health Evidence Guideline Quality Checklist (iCAHE),^75^ and use a yes/no rating system for each item, rather than a ten-point rating. This was done for ease of stakeholder use, and for ease of interpretation and scoring. See the Appendix D for the adapted and scored stakeholder checklist.

Given that the tool was modified, the results of the checklist were analyzed qualitatively rather than quantitatively. All four stakeholders provided positive feedback across the 23 items in the form of their yes/no ratings. Three out of the four stakeholders rated all 23 items positively.

One out of four stakeholders rated one item negatively, indicating that a procedure for updating the guidelines was not provided. This was the only negative rating on an item. As a result, the authors attempted to elaborate on the recommendation updating process in the final draft of the manuscript. In addition, subjective feedback from the stakeholders indicated that the recommendations were “very readable” and a “wonderful resource for clinicians and researchers alike.” Regarding next steps, one clinical stakeholder indicated that the process for updating the recommendations should be better outlined and more robust. However, given that this recommendations paper is one of the first of its kind in the field of educational speech-language pathology, there is limited consensus or documented procedure for how to update recommendations in a practice guideline of this type. In order to provide researchers and clinicians with up-to-date recommendations, these guidelines will be continuously updated as the body of literature progresses (see Recommendations Expiration and Updating section).

## Results

### Recommendations for Virtual Administration of Standardized Assessments

#### 1. CANDIDACY FOR PARTICIPATION IN VIRTUAL ASSESSMENTS

Participants and/or Clinicians *Must:*

- Participants must have access to a device connected to the internet;
- Participants must have a reliable and strong internet connection;
- Special accommodation must be made for participants who have significant behavioural, attentional, or cognitive difficulties. Clinicians must consider ahead of time how they will adapt the virtual testing process to support the participant. This could include:

- Having a caregiver present to ensure the child stays in the assessment session or to provide tangible reinforcements (snacks, toys etc.)
- Scheduling frequent mental and physical breaks
- Providing tailored reinforcement that interests and motivates the child on a regular schedule.
- Completing the session over several time periods to ensure the child is consistently performing their best
- Depending on the child’s abilities and needs, consider whether standardized assessment is appropriate
- Special accommodation must be made for participants who are hard of hearing or who have vision impairment. This could include:
- Specialized headphones that function with assessment equipment.
- The presence of a caregiver or adult who can troubleshoot audiology equipment like cochlear implants or hearing aids.
- Software that allows for enlarged images or coloured overlays.
- Depending on the child’s abilities and needs, consider whether standardized assessment is appropriate

Participants and/or Clinicians *Should*:

- Participants should have previous exposure to computers and have basic computer literacy skills;
- Participants should have desk readiness and the ability to sit and attend to a computer session;
- Consideration should be given for how different age groups may be more or less suitable for virtual standardized assessment;

- `o` Preschool-aged children can be more challenging to engage virtually, specifically those who have limited computer or desk experience.

##### 2. COMMUNICATION AND COLLABORATION WITH CAREGIVERS

Clinicians ***Must:***

- Obtain informed consent prior to the assessment and thoroughly explain all aspects included in standardized assessments. Provide the opportunity for caregivers to ask questions. This must also include informed consent to communicate via email and to audio record the assessment session;
- Obtain a completed background information form and questionnaire prior to starting the virtual assessment. Consider written or oral formats based on individual caregiver preference and ability.

Clinicians ***Should*:**

- Be flexible and provide caregivers choices. This may include completing assessment in chunks or staggered over multiple meetings to accommodate the participant;
- Consider meeting virtually or by phone with caregivers prior to the assessment to discuss the following:

- Determine what device the child will be using to complete the assessment. The clinician may recommend that a desktop or laptop computer is preferred
- to ensure the participant is seated in front of it at a table rather than lying on a couch or walking about the room.
- Remind caregivers about charging or plugging in portable devices to avoid loss of connection mid-assessment.
- Determine the location that the child will complete the assessment. Clinicians should emphasize that a quiet, private space, with adequate lighting and minimal distractions is preferred.
- Identify the interests of the participant to determine reinforcement and rewards if deemed necessary.
- Determine whether the caregiver will be present during the virtual assessment.
- If the caregiver attends the session, further directions should be provided regarding their positioning, ideally behind the participant in view of the camera.
- Outline rules for standardized assessments for caregivers. This should include information such as no repetition of instructions, no providing the participants with hints and no additional encouragement from the caregivers.

Additional Considerations:

- Consider reminding parents of upcoming assessments one week ahead of time, via email or phone call;
- Consider how caregiver presence during the assessment can be helpful for the management of behaviours, manipulation of testing materials on screen and provision of reinforcement, providing the caregiver is clear on standardized assessment rules;
- Consider how interpreting body language and nonverbal communication can be difficult through virtual assessment. Clinicians should strive to be clear and explicit in their communication with caregivers and participants.

#### 3. TECHNOLOGY AND EQUIPMENT

Clinicians ***Must:***

- Have an appropriate device, that is fully charged and equipped with a functional microphone and camera;
- Have a secure and stable internet connection;
- Have access to a software platform that allows for synchronous video and audio (i.e., Zoom, Skype, Google Meet, Microsoft Teams);
- Have proficiency using the necessary hardware and software;
- Check with individual test makers that versions of standardized assessments are valid for virtual use, and are able to be used through public-facing screen sharing;

- Many test makers of common S-LP assessment tools (Pearson and Pro-Ed) do not allow for public-facing screen sharing of their tests;
- Clinicians therefore must use screen-mirroring with a document camera to remotely share visuals of tests with clients;
- This requires clinicians to have at least two devices, one for the video conference platform and another (tablet or phone) to use as a document camera to capture images to be mirrored and shared from their device;
- If the clinician is using two Apple devices, screen mirroring can occur automatically, if the two devices are not Apple, additional software must be downloaded on each device to allow for this;
- When clinicians are using screen mirroring of these standardized testing measures, recording on the part of the client or clinician is not permitted;
- New clinicians or agencies who are purchasing new standardized assessments may wish to consider purchasing online versions that can be used virtually and in person.

Clinicians ***Should*:**

- Have and use a headset with microphone for increased speech clarity;
- Review the variety of software options available and test each version to see which best suits their needs (Zoom, Google Meet, Microsoft Teams, Skype);
- Consider if the software allows for screen sharing, shared mouse control, annotation, stamping or screen drawing, and continuous video feed of client when sharing a screen;
- Ensure caregivers and participants have familiarity with and access to the chosen software;
- Ensure that images on shared or mirrored screens are as close to 9 inches in size as possible, to maintain test standardization; as per test maker recommendations.^76^

Additional Considerations

- Consider using two devices, one for the manipulation of testing materials and one for video feed from the participant

#### 4. ADMINISTRATION OF STANDARDIZED ASSESSMENTS

Clinicians ***Must:***

- Follow standardized assessment rules and instructions to the best of their ability;
- Report and document any changes made to the standardization procedure, when following standardized assessment rules is not possible;

- For example, if a clinician must repeat a test item due to poor connection, this must be documented as a change in administration in their report.
- Ensure an appropriate testing environment setup for both the clinician and the participant;

- Ensure the participant is seated at a desk or table in a back-supported chair, facing the screen directly head-on.
- When possible, ensure the participant is using either a laptop or desktop rather than a tablet or phone screen.
- Ensure both clinician and participant are in a quiet, private space (ideally room with closed door), with minimal distractions (no toys or busy backgrounds), adequate lighting (front-lit, natural light) and minimal background noise, with strong internet connection (use WIFI booster as needed).
- Ensure their technological set-up allows them to see their materials and the participant.
- Coordinate with caregivers ahead of time if tests require manipulatives to ensure appropriate replacements are available at the time of the test;

- If manipulatives are not available in the home, then pictorial stimuli can be provided as an alternative, this must be documented in the reporting.
- Request caregiver permission and consent to audio record components of the session.

- Recorded sessions can allow for verification of responses to questions later and to allow clinician to focus on administering test rather than scoring in the moment. (Test makers only permit session recording when testing visuals are not being shared on mirrored);
- Determine how to modify tests of receptive language that require touching or point to an item, or tests of written language that require a written response. Clinicians can consider the following ideas:

- If available and appropriate, consider using tests that evaluate receptive language without the need for interaction with an image. For example, tests that require a child to listen to a passage and answer questions.
- When screen sharing or mirroring, if the software allows for shared mouse control, allow the participant to hover their mouse over their answer.
- Alternatively, have them circle or stamp using the annotation feature. This requires pre-teaching of these skills and may affect test standardization.
- If screen sharing is not possible, consider modifying how participants provide their responses, by having them identify the number associated with their answer (e.g., 1, 2, 3 or 4 on the PPVT-5 ^77^), or using a colour overlay on the test easel to have them name the colour of their chosen picture. This also affects test standardization.
- For tests of written expression or spelling that require a child to write using pencil and paper, the tester can consider requesting that the caregiver take photos of the final written output to share after the assessment is complete. If this is not possible, testers can also ask the child to hold up their work to the camera and take a screenshot of the image to save and evaluate.

Clinicians ***Should*:**

- Complete regular comprehension checks and listening checks with the participant. Clinicians can consider the following strategies:
- “Teach-Back” technique: after explaining an activity or providing instructions to the client, ask them to repeat what you just said or explain the activity instructions in their own words.
- “Repeat-after-me”: say a silly phrase/sentence and ask the participant to repeat to ensure they are paying attention.
- “Cueing” technique: Touching the eye to cue to look, touching the ear to cue to listen as required to prevent speaking out of turn or over one another.
- Complete regular checks to ensure screen sharing or mirroring is working;

- Ask the participant to describe what they see on their screen (e.g., “Tell me what kind of animal you see on the screen”, “Tell me what you see in the scene,” etc.)
- Prepare for implementation of reinforcement and breaks effectively. Clinicians should determine what types of reinforcement would be preferred and how frequently they might be required. Individual reinforcement can be provided between complete tests or subtests, but testers should check manuals prior to providing differential reinforcement after individual test items. Considerations include:

- Virtual Games

- https://www.thecolor.com/
- https://www.happyclicks.net/click-tap-games/index.php
- http://mrpotatohead.play.scriptmania.com
- https://www.silvergames.com/en/connect-4
- Movement Activities

- Jumping jacks, toe touches, belly breathing.
- Following along to a dance.
- Tangible Reinforcements

- Coordinate with caregivers to provide tangibles like food or stickers.
- Small stickers and bite-sized or single piece food items are preferable.
- Bathroom breaks

- Pre-arrange with the caregiver depending on age
- Visual Schedules

- Some participants may benefit from a schedule, visual or written, so they know what to expect in the testing session. This can be prepared as a slide show to share with participants ahead of time. Reinforcement and breaks can be built into this schedule.
- Clinicians should determine how to modify instructions where test items cannot be repeated if a participant missed instructions due to poor connectivity;

- For example, if connectivity interferes with the ability to hear the test item in a number repetition task, the clinician could elect to skip that item, or repeat the item. Any such modifications must be documented and reported.
- Clinicians should attempt to establish rapport with participants in a virtual assessment just as they would in an in-person assessment. Clinicians can consider the following ideas;
- Engage in a brief conversation about a preferred topic.
- Do a quick “full-body warmup” to get focused for the session (e.g., big stretch up, left, right, touch your toes, 3 deep breaths).
- Clinicians may wish to set up an initial meeting with caregivers and participants prior to assessment to get to know each other.

Additional Considerations

- Clinicians should be aware of whether conducting virtual administration of standardized assessment precludes children from accessing specific funding or resources. Certain regions may only grant access to funding or resources as a result of an in-person assessment and this should be considered prior to assessment.

#### 5. ETHICS, CONSENT, AND CONFIDENTIALITY

Clinicians ***Must:***

- Inform caregivers of additional risks associated with sharing information virtually and conducting assessments on virtual platforms. For S-LPs working in the public sector, organizations typically designate secure assessment platforms that meet national or provincial health-information privacy requirements. Clinicians, working in the private sector, must also ensure that selected assessment platform versions are PHIPA (Personal Health Information Protection Act) or HIPAA (Health Insurance Portability and Accountability Act)- compliant (see, Health Information Privacy, 2021 for an overview of HIPAA-compliant video conferencing platforms including Zoom for Healthcare and Skype for Business);^78^
- Obtain informed caregiver consent for all components of virtual session, just as they would for in person assessment. Ensure that caregivers have the opportunity to ask questions. Some additional consent considerations include:

- Risk and benefits.
- Communication via email.
- Use of virtual platforms.
- Confidentiality.
- Storage of information.
- Virtual dissemination of reports and documents.
- Consult their provincial, state-wide, or regional regulatory body’s position papers or practice guidelines and ensure they meet privacy law requirements and keep all personal health information as confidential and secure as possible;

- Review available virtual platforms, data transmission and data storage options to select the option that meets your regulatory body requirements.
- Use password protection, data encryption, two-factor authentication and a secure internet network when required.
- Ensure that they contact and consult with the provincial, state-wide, or regional body prior to providing services across regional boundaries and outside of their area of certification or licensure;
- Complete detailed documentation of all interactions in the same way they would for in person practice.
- Have clear rules about where and how long they will store and keep personal health information and inform caregivers of these rules;

#### 6. CONSIDERATIONS FOR BILINGUAL PARTICIPANTS

Clinicians ***Should*:**

- Match bilingual clinicians with bilingual participants who speak the same language;
- Encourage caregivers of bilingual children to be present to translate if general task directions or instructions relating to test requirements are unclear;
- When available, and with caregivers’ consent, use an interpreter to:

- Review information about assessment with caregiver and ensure comprehension and consent to proceed. If the caregiver is going to be present during the session, it is also imperative that they understand standardized testing rules (e.g., no prompting) prior to the start of the session.
- Translate any communication between a caregiver and participant, to ensure standardized assessment protocols are still being followed.

##### Recommendations Expiration and Updating

As previously stated, we consider these recommendations as a starting point for future guidelines development and research into online assessment by clinicians and researchers. These recommendations will be updated, as new themes emerge in future research investigating the in-the-field use of these recommendations by clinicians and researchers with typically and non-typically developing clinical populations. We anticipate that, as a result of the current COVID-19 pandemic, further research on tele-assessments will be conducted in the coming years. As such, we will conduct a preliminary literature search, within the next two years, to determine if there are sufficient tele-assessment studies, within the field of oral language and literacy research, to develop formalized clinical practice guidelines (CPGs) - based on a systematic literature review. Additional searches may also be conducted on a bi-yearly basis, based on a needs-based analysis of current literature. These updates will be developed by consulting the co-author leads in conjunction with a recommendation development group with a similar composition, as the current team, to ensure diversity of clinical and research experience conducting standardized assessments in virtual setting. These updates will be published to the following link: https://dataverse.scholarsportal.info/dataverse/virtualcare/ on the University of Toronto’s open access Dataverse.

## Discussion

In recent years, virtual care has become more widespread.^3^ Previously, the incorporation of virtual care was primarily driven by the desire to save time and better allocate resources and reach those in rural communities who lived without access to valuable medical and health services. However, widespread use of virtual care became necessary for clinicians and researchers alike with the outbreak of the COVID-19 pandemic and work-from-home orders, while guidelines to support virtual clinical and/or research decision-making are scarce. The current paper offers practical recommendations for virtual administration of oral language and literacy assessments to be used by clinicians and researchers.

Through the virtual administration of standardized tests, and in the development of these recommendations, group members in this study identified benefits and challenges associated with virtual assessment. Regarding the challenges, several group members referred to poor internet quality and its effect on their ability to administer certain tests. In particular, tests that include a timing component, like the digit, letter, colour and object rapid automatic naming subtests in the CTOPP-2 ^39^, or tests that require careful attention to specific sounds, like the repetition of non-words in the CTOPP-2 ^39^ were most affected by poor internet quality. Tests that require pictorial or word stimuli like the word and non-word reading tests and the EVT-2 ^37^ were impacted by the size of device that the participant was using to view the stimuli. In our experience, some families did not have access to computers, tablets, or monitors, and attempted to participate in the assessment using their phone. Group members also reported that some families struggled to access assessment meeting links, particularly those who did not have experience navigating and using virtual communication software. See Supplemental File 1 for interview analysis notes from authors 1, 3 and 4.

These issues are of particular concern for lower socio-economic status (SES) families who may lack the necessary internet access, software, or hardware.^79, 80^ Even in developed countries, such as the United States, 17% of children have no access to a laptop or desktop computer.^81, 82^ Furthermore, children belonging to lower SES households and marginalized Black, Indigenous and People of Colour communities are more likely to have limited or no access to a computer and/or internet connection.^81^ In Canada, children from lower income households are less likely to have access to their own internet-enabled device as compared to higher income households; further, as compared to higher income households children from lower income households are also more likely to *only* access the internet through mobile devices rather than personal computers, which may influence assessment quality.^80^ The first author experienced this firsthand, while assessing children virtually as a clinician at a school board. Children from low SES neighborhoods were less likely to have a computer at home. They often used school-issued devices (in this case, Chromebooks), or when those devices were not working or available, their caregivers’ phones. On several occasions, the school did not have sufficient devices available for those who required them. Consequently, these students were not able to participate in assessment with the school S-LP, and importantly also not able to attend their virtual daily classes with their peers and educators, placing them at greater risk of falling behind. In addition, the school-issued devices did not allow students and their families to easily join meetings on the platform that the S-LPs were instructed to use (Microsoft Teams) as they were designed for classroom use and the Google Meet platform. This was problematic, as Google Meet is not regarded to be as secure in terms of its’ personal health information protection properties as Microsoft Teams. This created additional barriers to participation in assessment for low SES families, which had to be resolved through meeting invitation sharing through the classroom teacher. These studies and clinical anecdotes suggest that the very populations who may have the greatest need to for oral language and literacy assessment, those who are marginalized, living in low SES or remote and rural communities, are often those who do not have access to the necessary tools to participate in such assessments. Prior research has demonstrated the negative influence of low SES on early oral language skills, such as vocabulary knowledge, and subsequent literacy development.^83, 84^ It is therefore important to continue providing in-person oral language assessments and intervention for children belonging to marginalized and low SES communities. Clinicians and researchers should keep in mind that virtual assessment might not be suitable to all segments of the populations, due to limitations associated with internet and computer access.

Virtual assessment may be particularly effective for bilingual children living in a country where the heritage language is not the dominant societal/educational language. Given the lack of practicing bilingual S-LPs across Canada and the US,^10, 11^ tele-assessment enables access to S- LPs and language interpreters speaking the same heritage language – without additional transportation or geographical barriers. For bilingual children, with limited or non-balanced proficiency in the societal/educational language, assessment across both heritage and societal languages can enable early identification of potential oral language and literacy issues as well as facilitate intervention in the dominant language.

This paper has identified that further research is needed to validate virtual administration of standardized oral language and literacy assessments commonly used by clinicians and researchers. In addition, due to the limited-permission nature of the No Objection orders, it is uncertain whether publishers will indefinitely allow clinicians to conduct virtual assessments using these tests, that were developed for in-person administration. These restrictions, along with a lack of standardized assessments validated for virtual use, limit the scope and type of assessments clinicians and researchers are able to conduct. For example, a commonly used tool for assessing phonological processing, the CTOPP-2 ^39^, has not been validated for virtual use. As a result, clinicians and researchers may be required to administer non-standardized assessments, or components of validated standardized measures, such as the Kaufman Test of Educational Assessment-3 ^85^, which require additional training and may not be as readily available in their place of practice. This paper has identified that further research is needed to validate virtual administration of standardized oral language and literacy assessments commonly used by clinicians and researchers, such as the CTOPP-2 ^39^, EVT-3 ^63^, and PPVT-5 ^77^, among others. Virtual assessments can also pose additional challenges specific to researchers. Virtual experiments may facilitate a larger sample size by limiting potential geographical location or transportation-related participation barriers. However, heterogeneity in terms of the type and quality of internet-enabled devices accessible to the child, such as whether the child completes the assessment via a limited-function and smaller mobile device as compared to a personal computer, may limit generalizability of research findings to children across diverse SES groups. Despite potential research-related challenges, virtual assessments can facilitate global cross-cultural speech-language research collaborations, while limiting potential cost and location-related assessment or research participation barriers.

## Limitations

The current recommendations are an initial step toward the development of evidence-based virtual assessment guidelines. These recommendations were developed based on the experience of 12 individuals using three standardized measures with a select population. We are cognizant that feedback from four stakeholders on an adapted checklist cannot be generalized to indicate that these recommendations would be rated positively by all clinicians and researchers working with standardized assessments in a virtual setting. Future iterations of these recommendations, and other more formalized Clinical Practice Guidelines, should be reviewed and rated by clinicians and researchers working in a variety of settings and with diverse experiences to ensure their usefulness, readability and to determine their quality. It is important to note, that all members and stakeholders who participated in this study had an interest in developing recommendations for virtual standardized assessments, which may have influenced the outcomes. This could theoretically be avoided in the future, through inclusion of members who have varied opinions on implementation of virtual practice. Ideally, reviewer and stakeholder groups should include clinicians and researchers with mixed opinions on virtual care, some with ample experience, as well as those who are new to this service delivery model. This would provide the opportunity to explore whether a) the recommendations are aligned with what experienced clinicians and researchers already do in their practice, and b) if the recommendations are useful for new clinicians and researchers in establishing good practices for virtual standardized assessment of oral language and literacy tests. Finally, we recognize the limitations of using a deductive analysis to evaluate the interviews in this study. Though the authors attempted to mitigate some of the potential bias by including an “*other”* section to provide an opportunity to address points that did not align with a given theme, it is possible that different responses might have been obtained from members, had the interviews not been structured in this format. Ultimately, this approach was chosen as it is less time-consuming than conducting an inductive thematic analysis, and the authors were motivated to share recommendations to their clinical and research colleagues in a timely manner for use during the COVID-19 pandemic.

## Future Directions

As additional research in the field of virtual language and literacy assessment becomes available, more formalized Clinical Practice Guidelines should be developed, that systematically evaluate and incorporate these studies into their recommendations. Beyond virtual care, there is also more generally a lack of Clinical Practice Guidelines available in the field of developmental speech-language pathology. Currently, those working in the medical sector, have access to select Clinical Practice Guidelines to inform certain aspects of their practice. Specifically, there are formal guidelines available for instrumental assessment of voice,^86^ and acquired velopharyngeal dysfunction.^87^ However, those seeking guidelines pertaining to intervention in the areas of speech, language, and literacy development, either in person or in a virtual model, may have to rely on general position papers from regulatory bodies, which often do not provide concrete practice recommendations, or individual research studies, which are often based on specific populations, limiting practice generalizability. Those working as clinicians and as researchers in the field of Speech-Language Pathology would benefit from the development and dissemination of additional Clinical Practice Guidelines addressing topics from developmental and medical sectors of the field. Consequently, future research should endeavour to evaluate the virtual administration of these types of assessments with children and adolescents with mild-to-severe speech and language difficulties and should expand the areas of oral language and literacy that are evaluated beyond expressive vocabulary, phonological awareness and word and non-word reading. Additionally, further research should consider the effect of child attention, caregiver involvement and access to necessary technology as a factor of SES on the virtual administration of such tests, and others.

It should also be noted that the majority of available standardized tools were developed for in person-use with monolingual-English speaking children. These standardized norm-referenced assessments are a valuable tool but are not the singular method of evaluating the language and literacy skills of school-aged children, particularly for children from linguistically and culturally diverse backgrounds whose English language skills often differ from their monolingual English peers. It will be critical for future research to investigate the validity of and provide guidelines for the administration of other non-standardized and informal evidence-based assessments, such as language sampling analysis and dynamic assessment in a virtual setting, with both monolingual and multilingual children.^88, 89^ As the body of research on virtual care continues to grow, researchers and clinicians must continue to collaborate on such studies to ensure that future guidelines are useful, realistic, and helpful for all.

## Conclusions

These recommendations cannot act as a basis for conclusive guidelines for all virtual clinical scenarios involving assessment of oral language and literacy skills. However, these findings provide a foundation for clinicians and researchers who are embarking on virtual assessment and who are seeking practical and useful suggestions to guide their practice and studies. They also provide a framework for future research into the feasibility and execution of online standardized assessments, as well as developing additional or revised guidelines.

An unexpected finding from this study and the development of these recommendations is that caregivers played a larger-than-expected role in the administration of the assessment tasks. Caregiver participation was critical in the management of behaviours, provision of reinforcement if required, translation of child responses from other languages, provision of interpretive feedback for the clinician or researcher as needed, as well as trouble-shooting any technical difficulties that arose during administration, particularly for younger participants. The caregiver was also required to assist if manipulatives were needed to complete tasks. Consequently, it may be valuable for future studies to consider whether there is an effect of caregiver participation on assessment results, as their involvement during the assessment may have unintended but significant effects on clients’ outcome. This notion should be further explored using a systematic approach, which also examines guidelines and recommended practices for parents involved in virtual assessments.

Secondly, the establishment of these recommendations has also illustrated that virtual assessment is a new skill to be learned by clinicians and researchers, similar to how standardized assessment is a new skill to be learned when clinicians are beginning their practice. Consistent, deliberate practice should be prioritized by those administering the tests to learn the nuances pertinent to administering the assessment virtually, as opposed to in-person. In anticipation of these challenges, academic institutions have begun providing instruction to clinical and research graduate students in the field of speech-language pathology in all aspects of virtual care, including virtual assessment. It will be critical for this practice to continue, to ensure that their graduates are qualified to practice or conduct research in the post-COVID era where virtual care has become ubiquitous.

## Supporting information

CORE-Q Checklist

Supplemental File - Interview Data

## Data Availability

 For confidentiality purposes, individual data is not available. Information is available in aggregate format in the tables attached in the appendices.

## Group Members

*Dr. Monika Molnar, PhD

Assistant Professor, Speech-Language Pathology, Rehabilitation Sciences Institute, University of Toronto, Toronto, Ontario.

*Emily Wood, M.Sc.A, Reg. CASLPO-C,

Speech-Language Pathologist, Durham District School Board, Whitby, Ontario. Volunteer, Bilingual and Multilingual Development Lab, University of Toronto, Toronto, Ontario.

*Insiya Bhalloo, M.Sc. Candidate

c. Candidate, Department of Speech-Language Pathology, Rehabilitation Sciences Institute, University of Toronto, Toronto, Toronto, Ontario.

*Brittany McCaig, MH.Sc. Candidate

Speech-Language Pathology Student, Department of Speech-Language Pathology, Rehabilitation Sciences Institute, University of Toronto, Toronto, Ontario.

*Cristina Feraru, MH.Sc. Candidate

Speech-Language Pathology Student, Department of Speech-Language Pathology, Rehabilitation Sciences Institute, University of Toronto, Toronto, Ontario.

Lisa Virtue, Speech-Language Pathologist 1, Durham District School Board, Whitby, Ontario.

Kathi Loughran, Speech-Language Pathologist 2, Durham District School Board, Whitby, Ontario.

Somyah Al-Ees, Volunteer and Speech-Language Pathologist Bilingual and Multilingual Development Lab, University of Toronto, Toronto, Ontario.

Siobhan Galeazzi-Stirling, Research Assistant 1, Bilingual and Multilingual Development Lab, University of Toronto, Toronto, Ontario.

Claire Liu, Research Assistant 2 Bilingual and Multilingual Development Lab, and Speech- Language Pathology Student, Department of Speech-Language Pathology, Rehabilitation Sciences Institute, University of Toronto, Toronto, Ontario.

Alisha Suri, Research Assistant 3, Bilingual and Multilingual Development Lab, University of Toronto, Toronto, Ontario.

Nicole Boles, Volunteer Research Assistant 1, Bilingual and Multilingual Development Lab, University of Toronto, Toronto, Ontario

* Starred members are also authors of this paper

## Stakeholders

Miriam Punnoose, Speech-Language Pathologist 4, Durham District School Board, Whitby, Ontario.

Ashley Sankowski, Speech-Language Pathologist 5, Durham District School Board, Whitby, Ontario.

Deborah Weinstein, Speech-Language Pathologist 6, Durham District School Board, Whitby, Ontario.

Alyssa Kuiack, M.Cl.Sc./PhD Candidate, School of Communication Sciences and Disorders, Western University, London, Ontario.

## Acknowledgements

The authors would like to acknowledge and thank the twelve group members who participated in the development and rating of these recommendations, as well as the four stakeholders who provided valuable insight and feedback on this paper, prior to its’ submission for publication. We are grateful and appreciative of your contributions.

