## Appendices for "Towards Development of Guidelines for Virtual Administration of Standardized Language and Literacy Assessments: Considerations for Clinicians and Researchers"

### Appendix A

#### *Identification and Development of Current Recommendation Paper Themes*

Table 1A. Results of Preliminary Narrative Review Search for Research Articles

| Retained Research Articles | | | | |
|---|---|---|---|---|
| **Article Citation** | **Population** | **Assessment Tool** | **Focus** | **Keywords** |
| Waite MC, Theodoros, DG, Russell TG, et al. Internet-based telehealth assessment of language using the CELF–4. *Lang Speech Hear Serv Sch* 2010; 41:445-458. | 25 children ages 5-9<br><br>Diagnosis of Language Impairment by S-LP<br>OR<br>Identified with difficulties in language by parent or teacher | Clinical Evaluation of Language Fundamentals – 4th Edition | Determine the validity of an Internet-based telehealth system for assessing childhood language disorders | Internet-based Telehealth<br>Telepractice<br>Standardized Language Assessment |
| Hodge MA, Sutherland R, Jeng K et al. Literacy Assessment Via Telepractice Is Comparable to Face-to-Face Assessment in Children with Reading Difficulties Living in Rural Australia. *Telemed J E Health* 2019; 25:279–287. | 37 children ages 8-12<br><br>Diagnosed with Specific Learning Disorder with impairment in reading | Woodcock Reading Mastery Test-III<br><br>Test of Word Reading Efficiency- 2nd Edition<br><br>MultiLit Sight Words Test<br><br>MultiLit Word Attack Test<br><br>Dalwood Spelling Test | Determine the feasibility and reliability of telepractice assessments for literacy assessments in children with reading difficulties | Telemedicine<br>Behavioural Health<br>Education<br>Telehealth<br>Pediatrics |
| Raman N, Nagarajan R, Venkatesh L et al. School-based language screening among primary school children using telepractice: A feasibility study from India. *Int J Speech Pathol* 2019; 21:425–434. | 32 children in grade 1 (M=6;3 years)<br><br>15 with concerns in hearing, speech, language or academics as reported by classroom teacher<br><br>17 with no specific concerns | Assessment of Language Development | Explore the feasibility of conducting school-based language screening using telepractice | Language<br>Telepractice<br>Screening |
| Sutherland R, Trembath D, Hodge A et al. Telehealth language assessments using consumer grade equipment in rural and urban settings: Feasible, reliable and well tolerated. *J Telemed Telecare* 2017; 23:106–115. | 23 children ages 8-12<br><br>History of reading difficulties and known or suspected language impairment | Clinical Evaluation of Language Fundamentals – 4th Edition | Examine the reliability and feasibility of conducting standardized language assessment with school-aged children via telehealth | Telehealth<br>Language Assessment<br>Rural<br>School-aged Children |

| | | | | |
|---|---|---|---|---|
| Waite MC, Theodoros DG, Russell, T.G et al. Assessment of children's literacy via an internet-based telehealth system. *Telemed J E Health* 2010;16: 564-575. | 20 children ages 8-13<br><br>Previous diagnosis with delays in reading and/or spelling by an SLP or school staff member | Queensland University Inventory of Literacy<br><br>South Australian Spelling Test<br><br>Neale Analysis of Reading Ability – 3rd Edition | Investigate the validity and reliability of an Internet-based videoconferencing system for assessment of children's literacy | Telehealth<br>Reading Disability<br>Speech-Language Pathology |

**Excluded Research Articles**

| Number of Unique Articles | Reason(s) for Exclusion of Articles |
|---|---|
| 129 | ▪ Not related to language or literacy (e.g. relating to speech, voice, fluency etc.)<br>▪ Not related to telepractice (e.g. relating to face-to-face practice)<br>▪ Not related to assessment, standardized or non-standardized (e.g. relating to intervention or treatment)<br>▪ Not conducted with a school-aged pediatric population (conducted with preschool-aged or adult populations) |

Table 1B. Results of Preliminary Narrative Review Search for Guideline and Recommendation Articles

**Guidelines Articles**

| **Article Citation** | **Guideline Qualifiers / Ratings** | **Article Themes** | **Corresponding Current Paper Themes** |
|---|---|---|---|
| Mashima PA and Doarn CR. Overview of telehealth activities in speechlanguage pathology. *Te lemed J E Health* 2008; 14:1101–1117. | No ratings or qualifiers used | Client Candidacy | Candidacy for Participation in Virtual Assessments |
| Richmond T, Peterson C, Cason J et al. American Telemedicine Association's principles for delivering telerehabilitation services. *Int J Telerabil* 2017; 9:63–68. | Shall – indicates a required action whenever feasible and practical<br><br>Should – indicates an optimal recommended action that is suitable<br><br>May – indicates additional points that may be considered to further optimize<br><br>Shall not – indicates this action is strongly advised against | Administrative Principles<br><br>Clinical Principles<br><br>Technical Principles<br><br>Ethical Principles | Administration of Standardized Assessments<br><br><br><br>Technology and Equipment<br><br>Ethics, Consent and Confidentiality |
| Brennan D, Tindall L, Theodoros D et al. A blueprint for telerehabilitation guideline s. *Int J Telerehabil* 2010; 2:31–34. | Shall-<br>[used for all guidelines, no definition provided] | Administrative<br><br>Clinical<br><br>Technical<br><br>Ethical | Administration of Standardized Assessments<br><br><br><br>Technology and Equipment<br><br>Ethics, Consent and Confidentiality |

Table 1C. Identification of Themes from Team Meetings

| Additional Sources for Development of Themes | | |
|---|---|---|
| **Source** | **Themes Identified Exclusively from Lab Meetings** | **Themes Identified in Lab Meetings and Existing Guidelines** |
| Lab Team Meetings | Communication and Collaboration with Caregivers<br>Considerations for Bilingual Children | Administration of Standardized Assessments<br>Technology and Equipment<br>Ethics, Consent and Confidentiality |

### Appendix B

***Guidelines for Virtual Assessment Lab Members Interview Script***

**A) Obtain participant consent for recording**

*Hello _____, thank you for agreeing to meet via Zoom to discuss advantages and challenges of virtual administration of standardized assessment protocols. This interview will focus on your online literacy assessment experience and it will be recorded. Only I will have access to this recording, it will be not shared with a third party. Are you OK with me recording this interview?*

**B) Proceed with following explanation and interview questions**

*This interview will last approximately 30 minutes and will be semi-structured. I will ask for your perspective on various aspects of virtual assessment. We will cover 6 different topics and there will be an opportunity to share 'other' insights at the end of this interview. Do you have any questions for me before we begin?*

**Sample / Candidacy for Participation**
**First, we will discuss candidacy for participation in virtual assessment. *Consider who is able to participate in virtual assessment and who is not and the reasons why this might be, e.g. SES, cognitive ability, access to technology etc.**
**What are the advantages?**
**What are the challenges?**

**Collaboration / Caregiver Communication**
**Next, we will discuss collaboration and communication with caregivers. * Consider communication with the guardian before, during and after the assessment in terms of setting up the assessment, the assessment itself, and the follow up on the assessment.**
**What are the advantages?**
**What are the challenges?**

**Technology / Equipment**
**Next, we will discuss the technological / equipment requirements. *Consider what software and hardware and testing materials the tester needs, what the child / guardian requires etc.**
**What are the advantages?**
**What are the challenges?**

**Clinical / Administration**
**Next, we will discuss the clinical administration of the standardized tests. *Consider the following: Test environment, Child interaction, Test administration, Test scoring.**
**What are the advantages?**
**What are the challenges?**

**Bilingual Children**
**Next, we will discuss special considerations for assessment of bilingual children.**
**What are the advantages?**
**What are the challenges?**

**Ethics and Confidentiality**
**Last, we will discuss ethics and confidentiality issues.**
**What are the advantages?**
**What are the challenges?**

**Other**
**Do you have any other advantages or challenges regarding virtual standardized assessment?**

**C) End Interview**

**Thank you for your participation in this interview. Please feel free to email me with other advantages or challenges should you have any.**

### Appendix C

***Group Member Recommendation Ratings – Round 1***

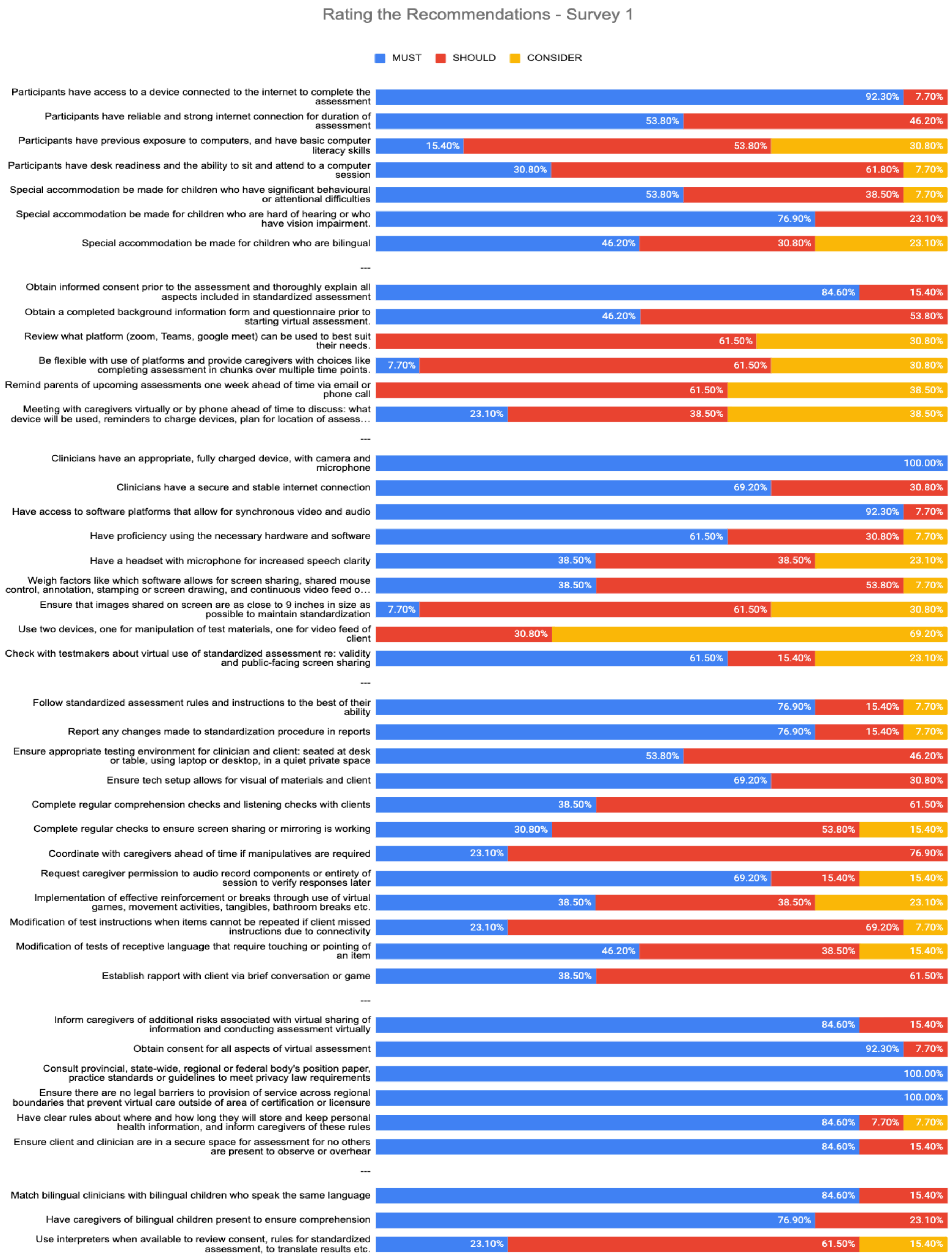

Rating the Recommendations - Survey 1
MUST
SHOULD
CONSIDER
Participants have access to a device connected to the internet to complete the assessment
92.30%
7.70%
Participants have reliable and strong internet connection for duration of assessment
53.80%
46.20%
Participants have previous exposure to computers, and have basic computer literacy skills
15.40%
53.80%
30.80%
Participants have desk readiness and the ability to sit and attend to a computer session
30.80%
61.80%
7.70%
Special accommodation be made for children who have significant behavioural or attentional difficulties
53.80%
38.50%
7.70%
Special accommodation be made for children who are hard of hearing or who have vision impairment.
76.90%
23.10%
Special accommodation be made for children who are bilingual
46.20%
30.80%
23.10%
---
Obtain informed consent prior to the assessment and thoroughly explain all aspects included in standardized assessment
84.60%
15.40%
Obtain a completed background information form and questionnaire prior to starting virtual assessment.
46.20%
53.80%
Review what platform (zoom, Teams, google meet) can be used to best suit their needs.
61.50%
30.80%
Be flexible with use of platforms and provide caregivers with choices like completing assessment in chunks over multiple time points.
7.70%
61.50%
30.80%
Remind parents of upcoming assessments one week ahead of time via email or phone call
61.50%
38.50%
Meeting with caregivers virtually or by phone ahead of time to discuss: what device will be used, reminders to charge devices, plan for location of assess…
23.10%
38.50%
38.50%
---
Clinicians have an appropriate, fully charged device, with camera and microphone
100.00%
Clinicians have a secure and stable internet connection
69.20%
30.80%
Have access to software platforms that allow for synchronous video and audio
92.30%
7.70%
Have proficiency using the necessary hardware and software
61.50%
30.80%
7.70%
Have a headset with microphone for increased speech clarity
38.50%
38.50%
23.10%
Weigh factors like which software allows for screen sharing, shared mouse control, annotation, stamping or screen drawing, and continuous video feed o…
38.50%
53.80%
7.70%
Ensure that images shared on screen are as close to 9 inches in size as possible to maintain standardization
7.70%
61.50%
30.80%
Use two devices, one for manipulation of test materials, one for video feed of client
30.80%
69.20%
Check with testmakers about virtual use of standardized assessment re: validity and public-facing screen sharing
61.50%
15.40%
23.10%
---
Follow standardized assessment rules and instructions to the best of their ability
76.90%
15.40%
7.70%
Report any changes made to standardization procedure in reports
76.90%
15.40%
7.70%
Ensure appropriate testing environment for clinician and client: seated at desk or table, using laptop or desktop, in a quiet private space
53.80%
46.20%
Ensure tech setup allows for visual of materials and client
69.20%
30.80%
Complete regular comprehension checks and listening checks with clients
38.50%
61.50%
Complete regular checks to ensure screen sharing or mirroring is working
30.80%
53.80%
15.40%
Coordinate with caregivers ahead of time if manipulatives are required
23.10%
76.90%
Request caregiver permission to audio record components or entirety of session to verify responses later
69.20%
15.40%
15.40%
Implementation of effective reinforcement or breaks through use of virtual games, movement activities, tangibles, bathroom breaks etc.
38.50%
38.50%
23.10%
Modification of test instructions when items cannot be repeated if client missed instructions due to connectivity
23.10%
69.20%
7.70%
Modification of tests of receptive language that require touching or pointing of an item
46.20%
38.50%
15.40%
Establish rapport with client via brief conversation or game
38.50%
61.50%
---
Inform caregivers of additional risks associated with virtual sharing of information and conducting assessment virtually
84.60%
15.40%
Obtain consent for all aspects of virtual assessment
92.30%
7.70%
Consult provincial, state-wide, regional or federal body's position paper, practice standards or guidelines to meet privacy law requirements
100.00%
Ensure there are no legal barriers to provision of service across regional boundaries that prevent virtual care outside of area of certification or licensure
100.00%
Have clear rules about where and how long they will store and keep personal health information, and inform caregivers of these rules
84.60%
7.70%
7.70%
Ensure client and clinician are in a secure space for assessment for no others are present to observe or overhear
84.60%
15.40%
---
Match bilingual clinicians with bilingual children who speak the same language
84.60%
15.40%
Have caregivers of bilingual children present to ensure comprehension
76.90%
23.10%
Use interpreters when available to review consent, rules for standardized assessment, to translate results etc.
23.10%
61.50%
15.40%

***Group Member Recommendation Ratings – Round 2***

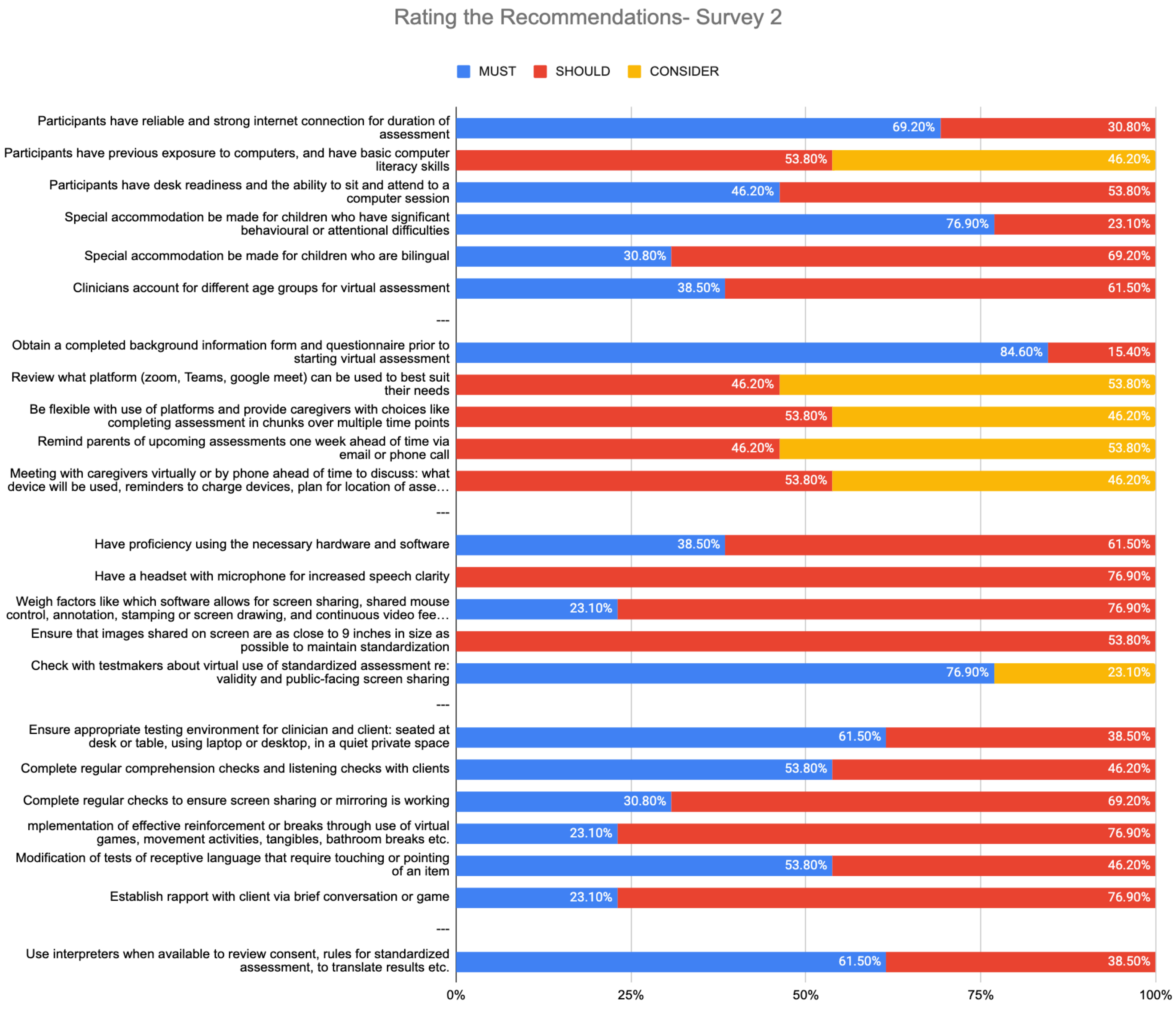


**Appendix D**

*Adapted AGREE-II Stakeholder Checklist and Results*

| Recommendations for Virtual Assessment Stakeholder Review | Yes | No |
|---|---|---|
| **Scope and Purpose** | | |
| Is the overall objective specifically described? | 100% | |
| The population for whom the guidelines apply is described | 100% | |
| The guideline development group includes individuals from all relevant professional groups | 100% | |
| The target users of the guidelines are clearly defined | 100% | |
| Is the target setting adequately described so that context for the guideline development is clearly understood? | 100% | |
| **Rigour of Development** | | |
| The criteria for selecting the evidence is clearly described | 100% | |
| The strengths and limitations of the body of evidence are clearly described | 100% | |
| The methods for formulating the recommendations are clearly described | 100% | |
| Could this intervention be replicated by other educators, researchers, or practitioners? | 100% | |
| A procedure for updating the guideline is provided | 75% | 25% |
| The guideline has been externally reviewed by stakeholders prior to publication | 100% | |
| Is any essential information missing? | 100% | |
| Are potential limitations reported and addressed? | 100% | |
| **Clarity of Presentation** | | |
| The recommendations are specific and unambiguous | 100% | |
| How would you describe the quality of the writing? [high quality / low quality] | 100% | |
| Are the guidelines user friendly and easy to access? | 100% | |
| **Applicability** | | |
| The guideline provides advice and / or examples of how the recommendations can be put into practice | 100% | |
| The potential resource implications of applying the recommendations have been considered | 100% | |
| Is the generalizability of guidelines addressed? | 100% | |
| Do the guidelines contribute concrete recommendations for future research (directed toward educators, researchers, or practitioners, as appropriate)? | 100% | |
| **Editorial Independence** | | |
| The authors declare that the research was conducted in the absence of any commercial or financial relationships that could be construed as a potential conflict of interest. | 100% | |
| **Overall Assessment** | | |
| How would you rate the quality of these guidelines? [high quality, low quality] | 100% | |
| Would you recommend these guidelines for use? | 100% | |
