## Supplemental File - Interview Data for "Towards Development of Guidelines for Virtual Administration of Standardized Language and Literacy Assessments: Considerations for Clinicians and Researchers"

**Supplemental File 1**

**Interview Analysis Notes**

| Theme | Advantages | Challenges |
|---|---|---|
| **1. Sample / Candidacy** | • Access to those who live remotely, further reach<br>• Able to see more students for assessment<br>• More accessible for older students<br>• Those without a car or who have multiple kids<br>• Those who cannot leave for medical issues<br>• Covid-19 restrictions<br>• Matching clinicians who speak certain languages with children regardless of locations<br>• Assessments can happen outside "working hours"<br>• Participants from a broader range of places<br>• Older children are familiar with zoom from using other online platforms for school can do with more independence<br>• Often better for students who might be anxious or nervous or take time to get used to a new environment<br>• Need to cater to individual child<br>• Can allow those with physical disabilities or without access to vehicles opportunity | • Accessibility for children with hearing impairment<br>• May exclude lower SES<br>• Difficult to assess with students you don't already know, not sure of their behaviour or levels or reinforcers<br>• Difficult for children with attention issues / distractible<br>• May be more difficult for non neurotypical students<br>• Requires access to internet<br>• Difficult to interpret accents or speech impairments at times<br>• Difficult with younger children, particularly 4yo<br>• School readiness and desk readiness<br>• Computer literacy both clinician, child and parent<br>• Those who have access to necessary technology may be self-selected of certain SES<br>• Difficulty for those with rural internet<br>• Further testing required for students with attention difficulties and their ability to participate in online Ax.<br>• Home environment and space in home, parent presence, sibling presence, need for devices |

**2. Caregiver Collaboration & Communication**

BEFORE

- Email is convenient and easy can be done at any time
- Easy to keep track of info exchange for note taking purposes
- Send an email to parent day before and day of to remind them
- Multiple methods of automated reminders available
- Likely best practice to meet with parents ahead of time to lay ground rules for assessment and explain their role

DURING

- Meet with parent for first five minutes at least to go over ground rules
- Parents can be helpful for managing behaviour and for reinforcing
- Also for managing manipulatives if needed for other tests
- Consider a video demo of dos and donts for parents participating in virtual standardized ax.
- Having parents there can be useful as they often know how best to motivate child

AFTER

BEFORE

- Don’t always get a chance to meet parent ahead of time and get to know them
- Parent should always inform child about what will be happening (assessment is happening)
- Emails are often ignored, phone calls are best

DURING

- Best to have parent there at the beginning for the first five minutes at least
- Parent may not always be able to be present at session (no adult in room)
- Better if parent is in the room, behind the child in view
- Can parent be there if assessment is to happen during working hours?
- Parent as mediator for assessment can be challenging (telling parent what to do) especially with different behaviour management styles or parents feel differently about how assessment is going
- Sometimes loss of body language and facial expressions can be challenging for managing behaviour can feel awkward or feel the need to be more direct
- Challenging to establish rapport with parent

| | | |
|---|---|---|
| | • Communication after assessment in the context of this study was limited due to research nature, but all felt it should be face to face | • Best to lay ground rules and be firm with parent about expectations as they may want their child to do better and may interfere with Ax.<br>• Loss of face to face component, consider a get to know you pre-assessment session to get to know parents, their style and the child, talks with teacher and people who know them before, long term consideration if virtual learning lasts (videos of child ahead of time?) |
| **3. Technology & Equipment Requirements** | SOFTWARE<br>• Zoom and other platforms are free<br>• Many different options, may be worthwhile to see what platforms child or parent is familiar with<br>EQUIPMENT<br>• Less volume of STUFF, easier to keep things online in one central place<br>• Many tests that don't have manipulatives can be easily transformed<br>• All the things you need are on your computer, less setup<br>• Clinicians will likely use paper protocols<br>• Some tests are (EVT) is easy to do<br>• Easier to score digital document | SOFTWARE & CONNECTIVITY<br>• ZOOM pro? Or other software<br>• Internet connection<br>• Need audio and video to match especially for tests where speech intelligibility is important<br>• Time restraints on zoom regular cost associated with premium<br>• Not able to share a zoom account among clinicians for overlapping sessions<br>• Those who are in remote areas who benefit from virtual services often have poor connectivity<br>• Connectivity issues can impact testing<br>HARDWARE<br>• Ask parents what devices they have ahead of time to prepare and ensure charged |

HARDWARE

- Minimal tech is essential, basic computer with webcam
- Can be done with one computer

OTHER

- Don't need a vehicle
- Can complete more assessment with reduced time needed in between
- Doing it from home is nice for both parties
- Can be done from "anywhere"

VIRTUAL PROTOCOLS RECS

- Virtual assessment protocols would be best if they were interactive, not just PDF
- one click to select correct/incorrect
- automatically plays audio files
- automatically tabulates scores
- automatically identified ceilings and basals
- able to click to navigate between tabs or tests
- Use of keyboard for scoring instead of mouse
- Cost issues of editable pdfs
- You also pay per paper protocol

- A device, or multilpe devices computer is best, ipad and phone less preferred for clinicians
- May be best to have access to headphones --> $$ awkward to ask parents to pay for additional items
- iPad is not best, camera wise or sound wise, also can be picked up and carried around
- Some use hard copies of assessments to score by hand
- Ensuring adequate battery
- Most clinicians felt two devices were necessary
- Definitely want a mouse
- Setting up guided access on iPad? Does that exist on computer?

EQUIPMENT

- Many assessments do not exist as online versions, must be adapted, is this allowed? Can testmakers do this for clincians in time
- When technology breaks down repetition is necessary but not always allowed on tests, does this need to be considered?
- Don't use word documents, they are not user friendly
- Rename CTOPP files
- Issue of scanning protocols from home
- Some tests (RAN with timing, audio, marking errors) are harder to do, consider audio recording

- Some tests do not have pdf versions, this can be done by clincians (EVT) but is time consuming up front
- Have to consider how to set up easel rather than share screen

COMPUTER LITERACY

- Parents need to have basic internet problem solving skills
-  Clinicians need more than computer basics, they need proficiency
- What has child used tech for in the past? Use a separate device from what they use for play
- Concerns are managing all of the tabs and still being able to see the child and maintain their attention not losing them

**4. Clinical**

TESTING

- Easier to score in a digital document
- Can be even faster for older children
- Keeping things formal and structured can be easier online
- Possibility for autoscoring documents
- Able to go back and listen to recording for things you might have missed, speech etc.
- Consider tech fatigue from online learning, when are the kids fresh and attentive

BREAKS + REINFORCEMENT

- Reinforcement / behaviour management – cannot just pull out a game from bag clinician has to be very present for each part, hesitant to give over control of game for break
- More challenging to manage behaviour
- Limited options
- Child may leave for break and not return, clinician must be part of break
- Challenging to have interactive breaks
- Sometimes hard to give adequate reinforcement virtually

and ready to look at computer screen?

ENVIRONMENT

- Testing can be done from anywhere, no limitations of vehicles etc, easier to find a space especially in busy schools where there is no space
- Multiple assessments can happen simultaneously, no clinic space limitations
- Safe comfortable space for children, less warmup test to environment and tester
- Easy for parents for access

CHILD INTERACTION

- Some feel easier or same as in person, just be animated
- Can be easier for children with anxiety or nerves about being in a clinic
- Most found that they were still able to make a connection with the child

**consider part of background information form being questions about devices, space, familiarity with tech, testing environment, space in home, sibling, what reinforcers are necessary --> what questions would we ask, could be done prior to assessment or in first five minutes

- Cant use video or ipad reinforcement, students may be satiated after sitting on computer
- Food or manipulatives requires set up
- Feedback is harder, no high fives or physical movement or tokens, harder to manage
- Determining appropriate incentives is important clinicians have to be creative and individualized

ENVIRONMENT

- Good lighting and quiet environment
- Need a separate quiet space for both clinician and child / size of house
- Cant control the environment the child is in and distractions
- Sibling presence can be challenging
- Cant always control for your and the childs environment, but this is true in person as well

TESTING

- Practice items are important, may want to add in more practice items to ensure comprehension, do comprehension or listening checks
- When children move out of frame or view cant bring them back, sometimes have to call parents to retrieve them
- Only allowed to play test items once, can’t repeat items,

considerations for online administration

- Keeping focus on you and not other fun things on computer
- What Is ideal time? Testing is long, multiple sessions may be required
- Managing CTOPP audio files and protocol and child interaction is hard
- Difficult to manage all pieces on screen for clinician as well as child interaction
- Hard to score in real time, may mean making on the fly clinical decisions more difficult
- Need better real time scoring options virtual protocols with autoplay of tracks
- Relabel audio files of ctopp so theyre not just numbers
- Screen sharing does not always comply
- Sometimes hard to interpret tests where very similar sounding phonemes (nonword repetition)
- How will we deal with tests with manipulaives
- Non response or poor internet connection
- Mute your mic when typing so they cant hear when you type notes?
- Can you repeat if child didn't hear due to connection?
- May not be able to see if child is writing things down
- Many felt scoring with pen and paper was easier

| | | |
|---|---|---|
| | | CHILD INTERACTION<br>• Some found it more challenging to build rapport<br>• Sometimes challenging to maintain while managing testing<br>• Where to look for eye contact, camera or at the child's face?<br>• Child cant see what you are scoring or writing which sometimes is nice, cant see correct / incorrect |
| **5. Special Considerations for Bilingual Children** | • Parents may be present to translate answers<br>• Many did not notice difference in assessing bilingual students | • Some children may reply in another language<br>• Can be challenging to understand accents<br>• Consider different reading L-R vs R-L when testing<br>• Differences in ability for receptive and expressive ability measured through PA skills<br>• Parents may prompt in a second language we don't understand |
| **6. Ethics & Confidentiality** | • Virtual or soft copy consent is easier to manage and not lose<br>• Should ensure parents know what they are consenting to<br>• No one knows the assessment is being done, compared to at school or in clinic, more private<br>• May be valuable to have audio and video but some parents might not want | • Added risks<br>• Spyware + confidentiality breach<br>• Security of zoom<br>• Concerns about clinicians using personal computers without encryption<br>• Getting consent forms at appropriate times<br>• Who has access to info, where will it be stored? Is it deleted from all locations |

| | | |
|---|---|---|
| | video (especially for those with hijabs) this cant be shut off on zoom<br>• Easier to book second appt on the spot | • Need a strict protocol for managing audio files<br>• Need clear informed consent about audio files and what happens with them<br>• Risks storing on computer<br>• Sometimes need to uploaded to drive<br>• Ensuring forms are signed and received ahead of time<br>• Ensure parents have informed consent about potential for interruptions and know about recording<br>• More challenging to continue to get informed consent to continue |
| **7. Other** | • The general trend has already been a move to online assessment, testmakers and clinicians, parents need to prepare for this<br>• Time saving on both ends | • Will need to be a new skill learned by children and adults<br>• A lot of time spent on ensuring the session runs smoothly, and less time spent on actually interacting with child<br>• These tests were done on typically developing students, what will happen with students who struggle |
